# The effect of mobility restrictions on the SARS-CoV-2 diffusion during the first wave: what are the impacts in Sweden, USA, France and Colombia

**DOI:** 10.1101/2021.02.01.21250935

**Authors:** Telle Olivier, Samuel Benkimoun, Richard Paul

## Abstract

Combined with sanitation and social distancing measures, control of human mobility has quickly been targeted as a major leverage to contain the spread of SARS-CoV-2 in a great majority of countries worldwide. The extent to which such measures were successful, however, is uncertain (Gibbs et al. 2020; Kraemer et al. 2020). Very few studies are quantifying the relation between mobility, lockdown strategies and the diffusion of the virus in different countries. Using the anonymised data collected by one of the major social media platforms (Facebook) combined with spatial and temporal Covid-19 data, the objective of this research is to understand how mobility patterns and SARS-CoV-2 diffusion during the first wave are connected in four different countries: the west coast of the USA, Colombia, Sweden and France. Our analyses suggest a relatively modest impact of lockdown on the spread of the virus at the national scale. Despite a varying impact of lockdown on mobility reduction in these countries (83% in France and Colombia, 55% in USA, 10% in Sweden), no country successfully implemented control measures to stem the spread of the virus. As observed in Hubei (Chinazzi et al. 2020), it is likely that the virus had already spread very widely prior to lockdown; the number of affected administrative units in all countries was already very high at the time of lockdown despite the low testing levels. The second conclusion is that the integration of mobility data considerably improved the epidemiological model (as revealed by the QAIC). If inter-individual contact is a fundamental element in the study of the spread of infectious diseases, it is also the case at the level of administrative units. However, this relational dimension is little understood beyond the individual scale mostly due to the lack of mobility data at this scale. Fortunately, these types of data are getting increasingly provided by social media or mobile operators, and they can be used to help administrations to observe changes in movement patterns and/or to better locate where to implement disease control measures such as vaccination (Pollina & Busvine 2020; Pullano et al. 2020; Romm et al. 2020).

## Introduction

The way that COVID-19 has affected human mobility all across the world is unprecedented. Combined with sanitation and social distancing measures, control of human mobility has quickly been targeted as a major leverage to contain the spread of the virus. One important feature of the response to the first wave was implementation of the sanitary cordon and placing draconian travel restrictions on the population to prevent the spread of the pathogen. The extent to which such measures were successful, however, is uncertain. Although human mobility data correlated well with the spatial spread of the pathogen in Hubei prior to the implementation of the sanitary cordon (Kraemer et al. 2020), the travel restrictions have been estimated to have delayed the epidemic progression throughout China by only 3-5 days (Gibbs et al. 2020). More globally, both the timing and extent of travel restrictions varied widely, providing an opportunity to perform a comparative analysis of the impact of lockdown on the spread of SARS-CoV-2. In our hyper-connected world, there is extensive use of mobile phones, internet providers and social media and there is currently much discussion on the use of such data to guide the public health response to the Covid-19 epidemic (Grantz et al. 2020). Within strict guidelines concerning personal privacy, several mobile phone operators, social network and internet service providers across the world have offered their data at an aggregated level to be able to visualise patterns of mobility and contact (Buckee et al. 2020; Pollina et al. 2020; Romm et al. 2020). In this study, we quantify mobility and changes thereof and assess the impact of lockdown measures in four different countries, Sweden, USA (West Coast), France and Colombia, which instigated differing levels of lockdown.

## Objective

The objective of this research is to understand how spatio-temporal mobility patterns and SARS-CoV-2 diffusion are connected in four different territories: the west coast of the USA, Colombia, Sweden and France. Using data from the Facebook Data for Good program, which provides data on users sharing their location with the application, we first aim to address the impact of lockdown strategies on mobility patterns between administrative geographical units of each country. Then we estimate the contribution of these mobilities to the diffusion of SARS-CoV-2 among administrative units within each territory. Analysing the relation between SARS-CoV-2 spatial diffusion and changes in mobility provides insight into the efficiency of different public health strategies across the four countries that implemented diverse strategies to tackle the diffusion of the disease.

## Methods

### Mobility and Covid-19 Data (Table 1)

To measure mobility patterns during the pandemic, we accessed Facebook mobile users’ data provided within the company’s Data for Good framework through a dedicated platform. With the most up-to-date information, these data help us understand dynamically where people are. Measured every eight hours (00:00AM, 8:00AM, 4:00PM GMT), they provide the number of people who moved from a given administrative unit to another between two time periods. In this article, mobility is understood under this specific definition. The panel of users hereby monitored is made of users over 18-year-old, who shared their location with the mobile application. As shown in Table 1, this represents between 5.8% and 11.1% of the adult population across the studied countries. The mode of detection is passive, fully anonymised and no individual information is provided. The data are only accessible in a spatially aggregated form. Covid-19 case data (Sweden, USA and Colombia) and positive test data (France) were retrieved from the Public Health websites of each country.

**Table 1.** Mobility and Covid 19 Metadata.

| Country | Covid-19 data source | Covid granularity | Facebook data granularity | Starting date for mobility collection | Number of the FB mobile app with shared location | Percentage FB over 18 years old | Official covid data website |
| --- | --- | --- | --- | --- | --- | --- | --- |
| France | Santé Publique France | Département | Département | 3rd March | 3 138 246 | 5.80% | https://www.santepubliquefrance.fr/ |
| USA | Centre for disease control | County | Pitney Bowes -scaled down to County | 11th March | 3 984 921 | 11.10% | https://www.cdc.gov/ |
| Colombia | Salud y protection social | Municipios | Municipios | 13th March | 2 666 887 | 7.40% | https://www.minsalud.gov.co/Paginas/default.aspx |
| Sweden | The Public Health Agency of Sweden | County | County | 14th April | 572 617. | 6.70% | https://www.folkhalsomyndigheten.se/the-public-health-agency-of-sweden/ |

To link mobility and SARS-CoV-2 diffusion we need to match mobility information at the lowest available case resolution provided by the official health agency from each country. Facebook movements from one administrative unit (France – Department; USA (West-Coast) – County; Colombia – Municipios; Sweden – County/Län) to another were summed by week for every unit. For Colombia and France, the resolution of mobilities and Covid 19 cases were matching, but for the USA and Sweden we needed to downscale the mobility to that between counties. Unfortunately, we could not access all the mobility data of Colombia as Facebook provided only users’ mobility for 80% of the territory. The USA were limited to the West Coast, corresponding to the states of California, Oregon, Washington and parts of Nevada, Arizona, Idaho, Montana and Utah. All mobility data were processed with R (spatial and temporal aggregation, compilation of all the movements in one file…) and then mapped with QGIS, an open-source Geographic Information System (QGIS; R core team 2020).

### Epidemiological analyses

The number of cases in the unit of origin was summed per week and multiplied by the summed Facebook movements from unit of origin to unit of destination for the same week. This thus generated a potential incoming force of infection (*FoI*) for every unit of destination based on every unit of origin number of cases and incoming mobility (see Figure 1). This hybrid indicator combines the mobility connectivity of a geographical unit with the epidemiological context in the related units. The summed values (N) for each unit of destination were then log(10)(N+1) transformed yielding the variable *log(FoI)*:

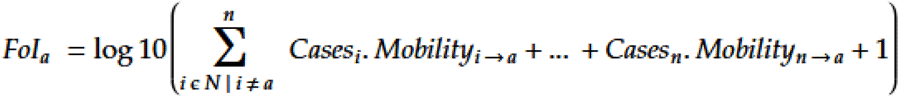

**Figure 1:**
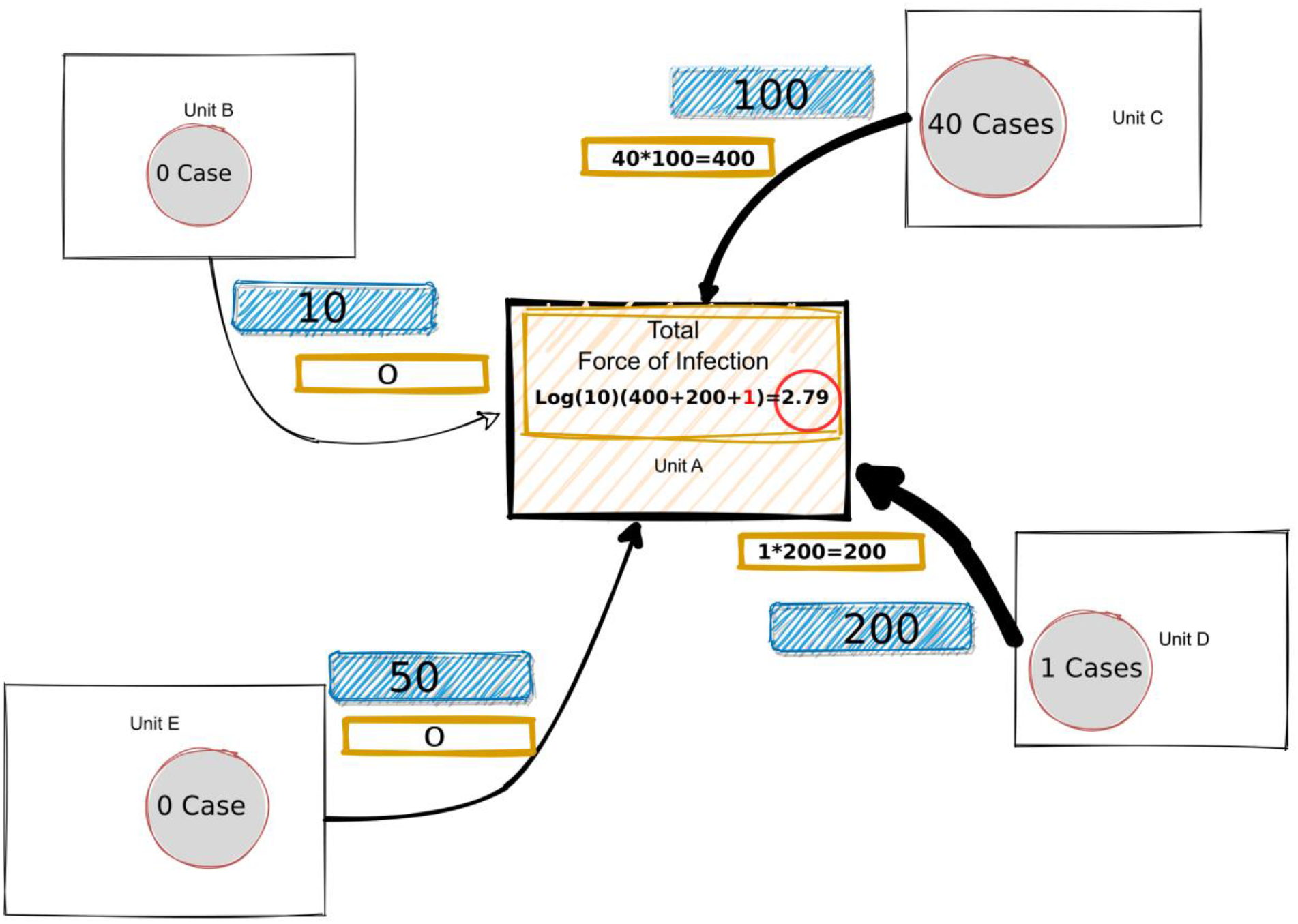
Theoretical model created to compute Force of Infection for unit A, with Facebook users mobility traveling from units B, C, D, E (in blue), number of Covid-19 registered in one unit (grey and red circle) and related FoI (yellow rectangle).

A Generalized linear model (GLM) with a logarithmic Quasi-Poisson distribution (recommended for count data with over-dispersion, Ver Hoef & Boveng, 2007) was then fitted to the number of cases per week per administrative unit as a response variable. As explanatory variables, we included the new number of cases registered in each unit during the previous week, and the *logFoI* from the previous week, plus their interaction term. We included the log(e) of each unit’s population as an offset value. The model was refitted at each time step (weekly) to give a dynamic perspective of the varying impact of the *FoI* through time, and from one country to another.

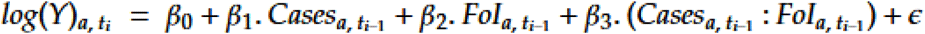

In order to evaluate the impact of the *log(FoI)* in the model, we computed the Quasi Akaike Information Criterion (QAIC) to assess the added value of increasing model complexity. The lower the value, the better the likelihood of the model is, given that adding a variable is penalised (here the penalty = +2 points for each added term). The value has no absolute meaning outside of the model’s own selection environment (i.e. no cross-country comparison), but serves as a comparator between different models fitted on the same dataset, to determine if an increase in the model complexity really improves the model fit. We then created box plots to display the diverse values of the QAIC from each week’s models. The QAIC function from the MuMIn *R* package was used (Barton & Barton, 2015). Then the *β* coefficients from the regression were examined. Since the model is based on a logarithmic distribution, the exponential of the coefficients is giving the multiplication term on the response variable by 1 unit increase in the explanatory variable. Applied to the *log(FoI)*, we then verified how influential this variable was on the number of cases of the following week. We also calculated the McFadden’s Pseudo-R_2_ (defined as 1−LLmod/LL0, where LLmod is the log likelihood value for the fitted model and LL0 is the log likelihood for the null model which includes only an intercept as predictor) to have an estimation of the goodness of fit of each country’s regression, and the p-values to assess statistical significance. All these steps were carried out in R.

### Concentration Index (Gini I)

The Gini coefficients of the countrywide distribution of cases (or infections) and mobilities were calculated to assess the extent of case and mobility clustering.

## Results

### Mobility pattern reduction and geography of mobilities

In Colombia, on March 20, the president announced a 19-day lockdown, starting on March 24. Before the lockdown, there were more than 3.5 million inter-municipios mobilities per week and 500,000 per day. These mobilities were mainly located around the geographical units containing the major cities of Bogota, Cali and Medellin, representing more than 65% of all inter-unit mobilities. The lockdown had a strong impact on inter-unit mobilities leading to an overall reduction of 78% between March 27 and April 2 (Figure 2). While Bogota, Cali and Medellin were well connected before the lockdown, Facebook data did not detect any mobilities among these cities in the post-lockdown period, and long distance mobilities (more than 50km) reduced by 93% (see collection of maps 1). At the local level, the reduction of mobilities around major municipios reduced by 70%. After April 10, even though cases continued to increase, inter-unit mobilities increased slowly but steadily to reach 68% of pre-lockdown level by mid-July. Overall, mobilities were highly concentrated to certain units of Colombia - the major metropolises of Colombia-before, during and after the lockdown, and the Gini index was approximately 0.95 at all times (Tables 2 a & b).

**Table 2a.**
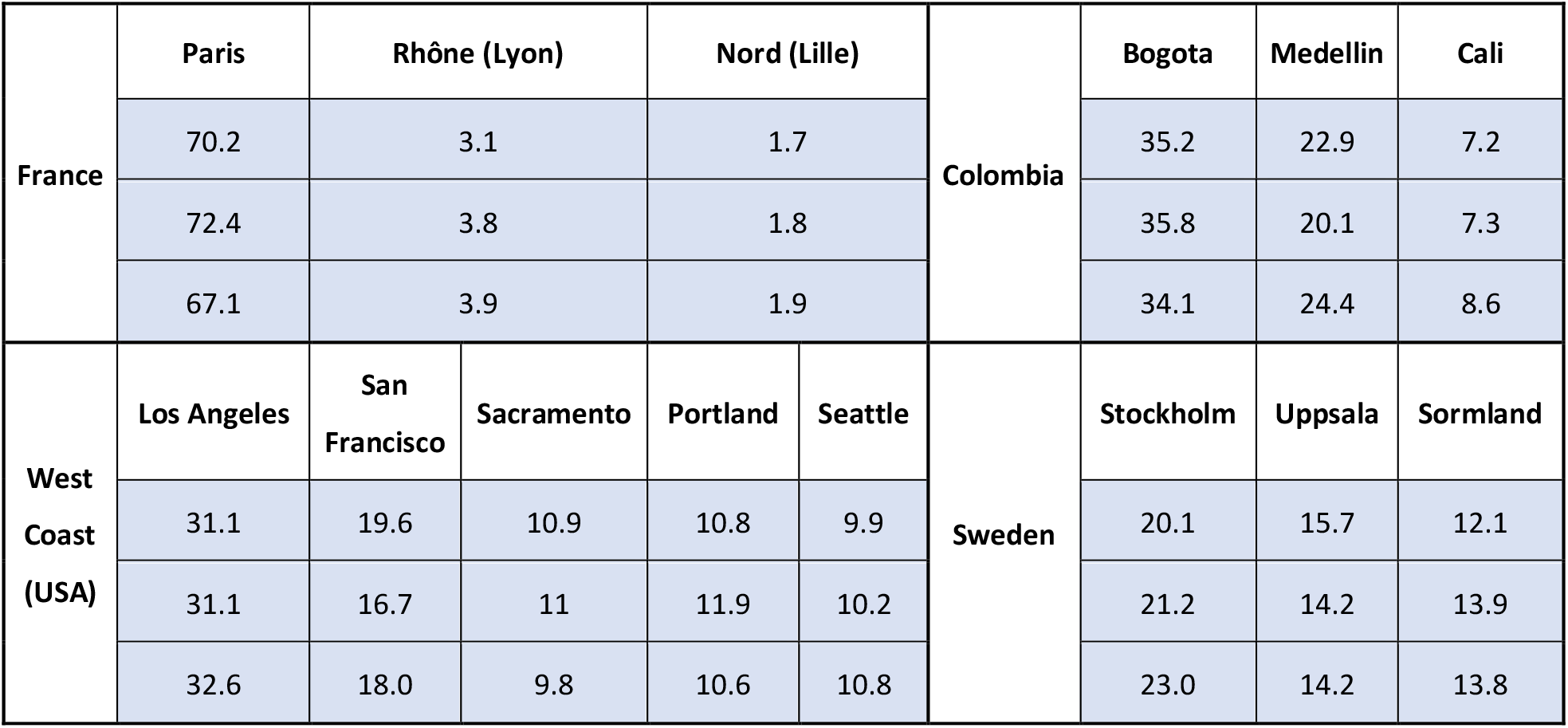
Percentage of inter-unit mobilities captured by different metropolitan areas.

**Table 2b.** Gini index related to mobility at different lockdown time-steps for the four countries.

|  | Colombia | France | West Coast | Sweden |
| --- | --- | --- | --- | --- |
| Gini Before lockdown (1st week of the data) | 0.95 | 0.76 | 0.77 | 0.76 |
| Gini Lock (1st week after the lockdown) | 0.95 | 0.79 | 0.77 | 0.60 |
| Gini last week of August | 0.96 | 0.74 | 0.72 | 0.59 |

**Figure 2.**
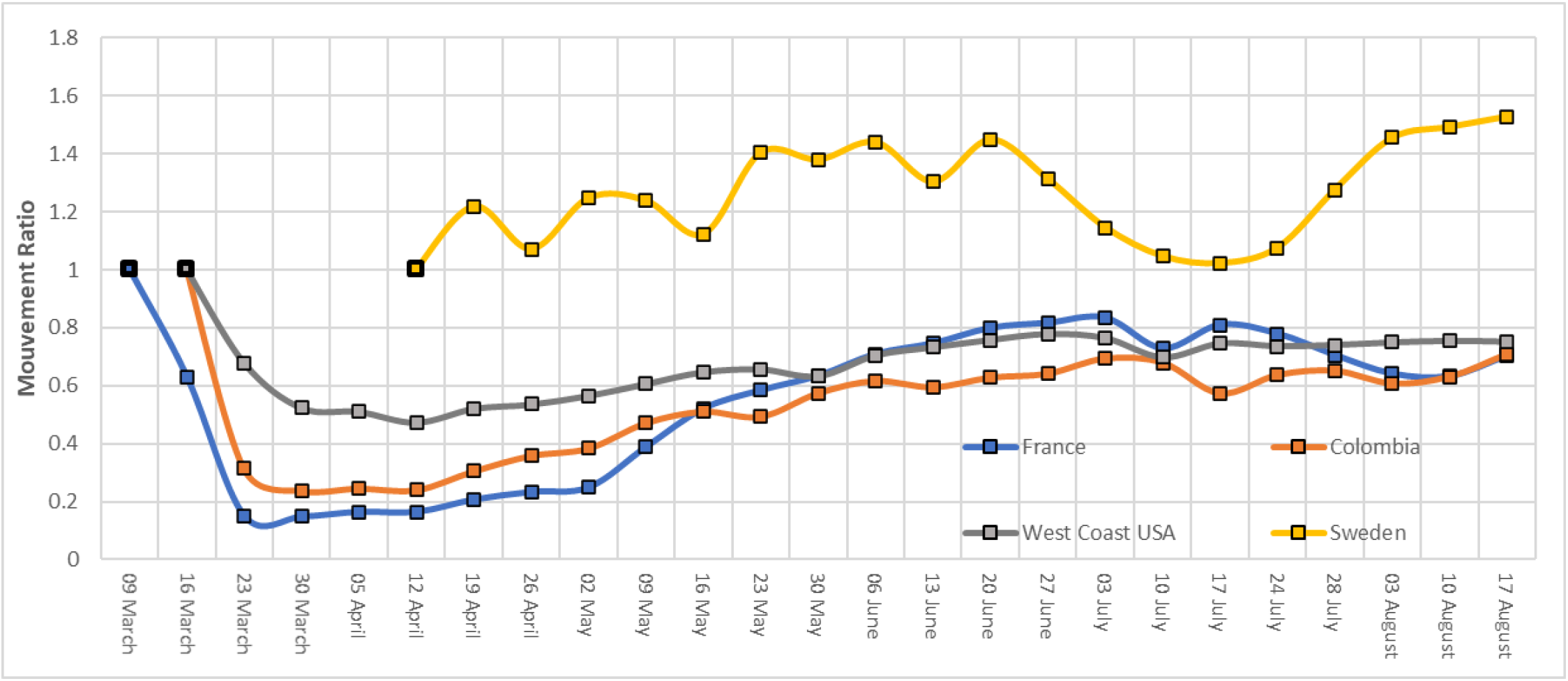
Evolution of inter-unit mobilities in the four countries. We use a ratio to compare the effect of lockdown in each country. The Black box for each country indicates the reference date for establishing the ratio for each country.

In France, lockdown measures were announced on March 16 (to be implemented the next day at 12 AM). The mobility among departments decreased by 82% during the peak of the lockdown before increasing slowly but steadily after the ease of the lockdown (Figure 2). The highest decrease in movement was observed at long distances during the peak of the lockdown, with no mobilities detected at distances >150km after March 27. As a consequence, the major cities were disconnected from each other (see collection of maps 2). As in Colombia, the majority of national movements before and during the lockdown were located in and adjacent to departments containing the major French cities. Mobilities to the departments of the Nord region (with Lille as a main metropolis), Bouches-du-Rhône (Marseille) and Rhône (Lyon) concentrated only 7% of overall inter-department mobilities over the period, whereas mobility between Paris and surrounding departments captured more than 66% of all inter-department mobilities (Table 2a). With a Gini index of 0.76 before the lockdown, incoming mobility concentration increased after the lockdown (G=0.79), to decrease again after the ease of the lockdown (G=0.74)(Table 2b).

In the United States, the sanitary measures taken were not centralized, unlike in other countries. This resulted in a very heterogeneous implementation of measures taken among states, counties and municipalities to counter the advance of SARS-CoV-2. For example, California was the first state to implement “Stay at home orders” on March 19, whereas these were not taken in Nevada and Arizona until April 6. Globally, inter-county mobilities remained relatively high, with 48% of inter-county mobilities maintained at peak lockdown (From April 6, graph 3), even if high distance mobility (>150km) decreased by 62% (Figure 2). During this lockdown period, main cities such as Los Angeles, San Francisco, Seattle, Sacramento and Portland remained highly connected to other more local counties, at an intermediary distance (between 20 to 150km) (see collection of maps 3). Metropolitan areas of each county captured more than 82% of inter-county mobility (Table 2a). As in France and Colombia, a small number of counties adjacent to the large cities concentrated the majority of the inter-county movements of the USA West Coast (G=0.77 before the lockdown) (Table 2b).

The Sweden scenario was exceptional since no travel restrictions were imposed at that time. The mobility data from April 5 showed a slight increase in inter-county mobilities from 13 April until 15 June (Figure 2). Overall, no significant reduction in mobility was observed at any time. Mobility data revealed less centralised movement patterns than observed in the other countries, since the most connected counties (Stockholm and Uppsala) concentrated only 25% of the incoming/outgoing population (Table 2a). This is translated into a Gini index that was the lowest of all the countries (G=0.59-0.76), and, despite no lockdown, became increasingly homogeneous over the same epidemic time period as the other countries (Table 2b).

### Epidemiological analyses

During the period March16 to August 31, the part of Colombia selected recorded 501,701 cases peaking in the week starting on July 17 with 53,829 cases (Figure 3). Santa Fe de Bogota was the most affected municipio with 246,395 cases registered by August 31, followed by Cali (57,640 cases). These two units recorded more than 60% of all cases registered in the studied area (Table 3a). The countrywide case distribution was highly concentrated before and during lockdown (G=0.98) and slowly decreased as the epidemic progressed and the virus spread nationally (G=0.87 in August) (Table 3b).

**Table 3a.** Percentage of cases captured by different metropolitan areas.

| France | Paris Metropole | Rhône (Lyon) |  | Nord (Lille) |  | Colombia | Bogota | Medellin | Cali |
| --- | --- | --- | --- | --- | --- | --- | --- | --- | --- |
|  | 59.3 | 5.1 |  | 0.3 |  |  | 46 | 15.2 | 14.8 |
|  | 46.1 | 3.2 |  | 0.8 |  |  | 60.1 | 8.2 | 17.5 |
|  | 33.3 | 6 |  | 2.8 |  |  | 45.2 | 15.3 | 7.11 |
| West Coast (USA) | Los Angeles | San Francisco | Sacramento | Portland | Seattle | Sweden | Stockholm | Uppsala | Sormland |
|  | 9.80 | 28 | 2.4 | 0.9 | 44.4 |  | 38.1 | 5.2 | 7.8 |
|  | 45.10 | 10.1 | 2.4 | 1.2 | 10.3 |  | 26.3 | 6.1 | 6.5 |
|  | 39.10 | 10.6 | 5.3 | 1.1 | 3.1 |  | 25.1 | 5.0 | 8.2 |

**Table 3b.** Gini index related to mobility at different time-steps and for the four countries.

|  | Colombia | France | West Coast | Sweden |
| --- | --- | --- | --- | --- |
| Gini Before lockdown (1st week of the data) | 0.98 | 0.78 | 0.91 | 0.57 |
| Gini Lock (1st week after the lockdown) | 0.97 | 0.70 | 0.85 | 0.5 |
| Gini last week of August | 0.87 | 0.66 | 0.72 | 0.59 |

**Figure 3.**
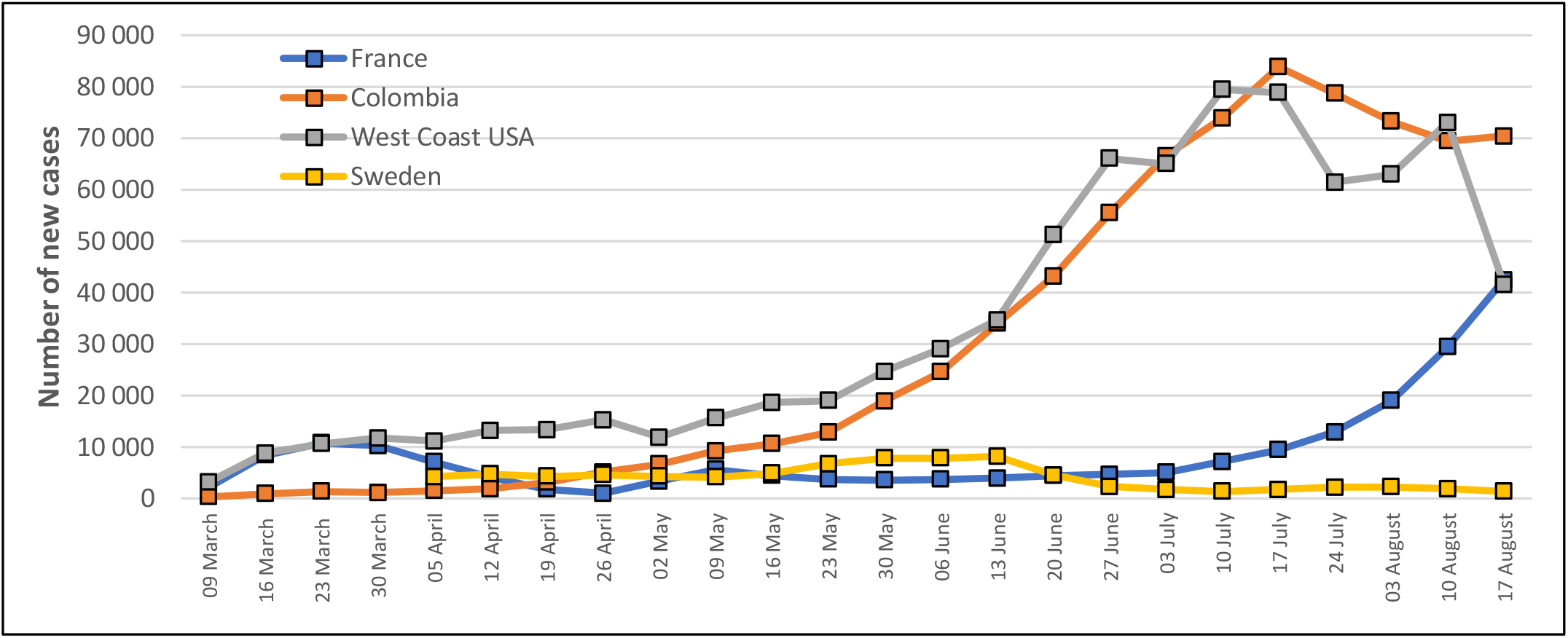
Weekly evolution of Covid 19 incident cases from March 9 until August 24

In France, from the period of March 5 until August 28, there were 197,550 positive tests (Figure 3). If the percentage of units detecting at least one case was higher than in Colombia when the lockdown was implemented (76%), a small number of units still captured the majority of cases (G=0.78) (Table 3b). The units concentrating most of the cases over the studied period were located around Paris (Ile de France), which concentrated from 59.3% to 33.3% of all Covid cases detected, followed by Lyon (3.1%-6%) and Lille (0.3%-2.8%)(Table 3a). As in Colombia, the Gini index tended to decrease during the epidemic (G=0.74 during the lockdown, G=0.66 in August)(Table 3b).

In USA, in the counties of the states of California, Idaho, Montana, Nevada, Oregon and Washington, from March 10 until August 31, there were 849,278 cases with a peak in the week starting on the August 3, with 80,721 cases (Figure 3). Once again, the majority of cases were concentrated in a small number of units before the lockdown (G=0.91) and the Gini index decreased steadily over the period to reach 0.72 in August (Table 3b).

In Sweden, the peak of the epidemic was reached by June 6, with more than 8,000 cases registered in a week (Figure 3). The nationwide case distribution was much less clustered in Sweden at all times and mirrored that of the population mobility distribution (Tables 2b and 3b).

### Force of Infection

In Colombia and France, the log(10) transformed Facebook weighted force of infection (FoI) explained a considerable amount of variation in weekly unit case number (20-50%, mean 41.2, SD 7.8), with only a small decrease in the weeks that followed the strictest lockdown period in both the countries (Supplementary Figure 1). The FoI explained far more of the variation than cases in the previous week in that unit (mean 17.4%, SD 7.6). There was a small but significant negative association of case number with the interaction term of previous week’s cases and FoI (% variation explained mean 7.2% SD 5.9).

In contrast to Colombia and France, in USA the majority of the variation in the current week’s case number was explained by the previous week’s case number in that county (mean 17.8% SD 15.8) and most notably during the first 12 weeks of the epidemic (until June 7)(Supplementary Figure 1). However, the FoI did explain a considerable amount of county case number variation (1-20%, mean 9.2% SD 6.4). Once again, there was a small but significant negative association of case number with the interaction term of previous week’s cases and FoI (% variation explained mean 15.0% SD 10.5). In Sweden, there was no significant association with FoI or with the interaction term on the current week’s case number.

As shown by the significantly decreasing Quasi Akaike Information Criteria, fitting the FoI, case number and their interaction largely improved the goodness of fit of the model for Colombia, France and USA. Globally, fitting the complete regression model improves the model performance as compared to fitting a model with only cases from the previous week (Fig. 4). There was a significant and consistent improvement of the likelihood with the full model for Colombia *(median[Intercept +Cases week-1]= 415 vs. median[I+C*_*-1*_*+FoI+(C*_*-1*_*\*FoI)=279)*, France *(mean[I+C*_*-1*_*]= 128 vs. median[I+C*_*-1*_*+FoI+(C*_*-1*_*\*FoI)=106)* and USA *(mean[I+C*_*-1*_*]= 129 vs. mean[I+C*_*-1*_*+FoI+(C*_*-1*_*\*FoI)=116)*. In the case of the USA, the interaction term played an important role since the QAIC only decreased after its addition, meaning the *log(FoI)* predictive power was significantly moderated by the number of cases. However, for Sweden there was no gain in increasing the complexity of the model *(mean[I+C*_*-1*_*]= 30 vs. mean[I+C*_*-1*_*+FoI+(C*_*-1*_*\*FoI)=31)*.

**Figure 4.**
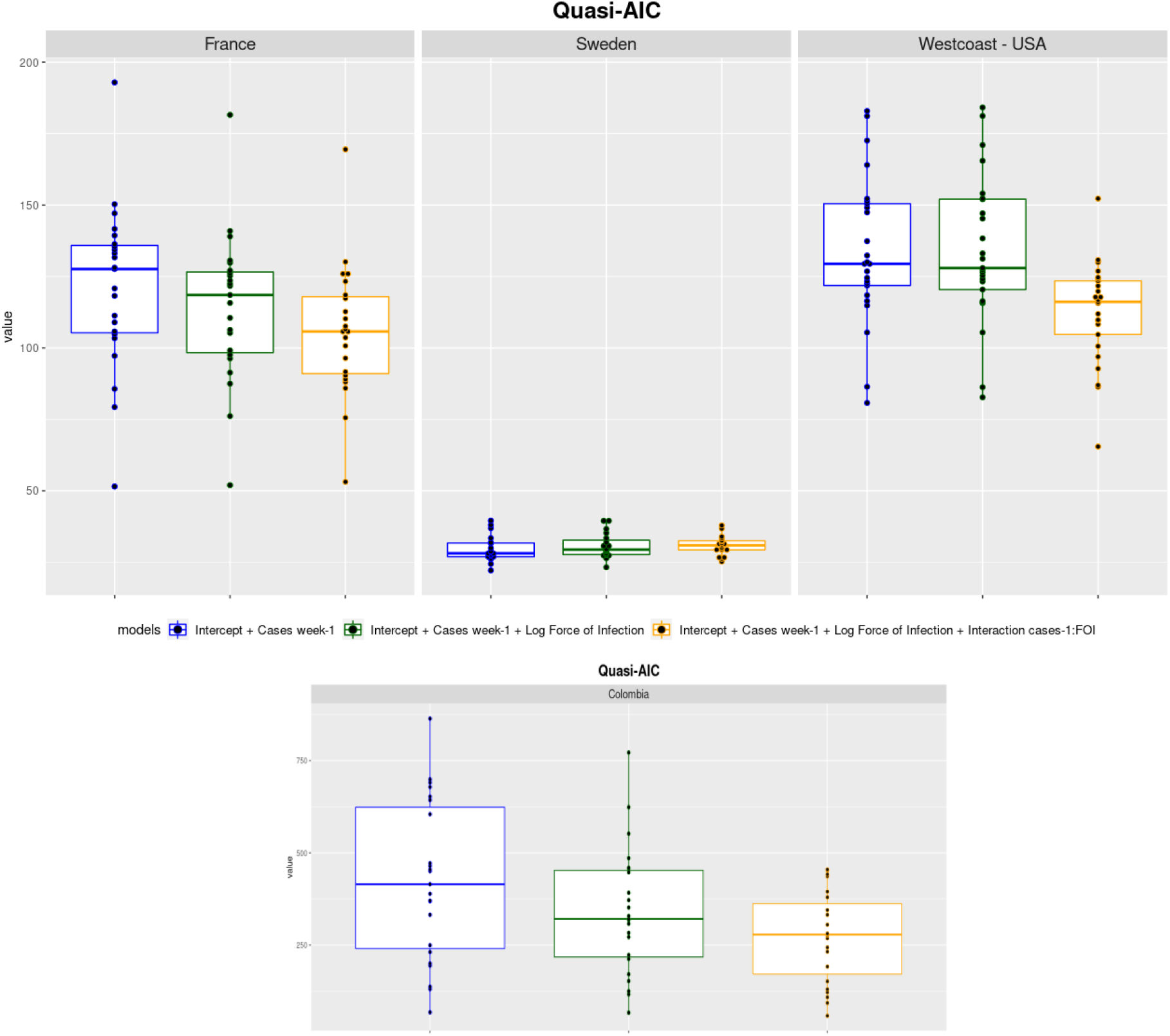
Boxplots of the QAIC values taken by each model while updating for each week.

To assess the exact strength of the nexus between this mobility driven variable (*log(FoI)*) and the number of cases per week (Figure 5), the exponential of the regression coefficients at each time step was calculated. This shows the size of the multiplicative term on the cases of the current week that an increase of 1 unit in the *log(FoI)* of the previous week would generate. For Colombia it was relatively constant, with an average of 1.30 over all the time period. In France, although more variable, the strength of the association was stronger, averaging at 1.47. For these two countries, we can see a common temporal tendency with a decrease of the FoI effect following the first lockdown. The effect then increases again from May onwards, probably because of the alleviation of the mobility restriction in both countries at that time. In the USA, the effect was very mild, with an average of 1.15 and peaking in June. By comparison the multiplicative term was only 1.18 on average for Sweden with a confidence interval that often overlapped with 1, meaning that the relationship between those two variables is not even clearly positive overall, despite two main peaks in the beginning of June and July.

**Figure 5.**
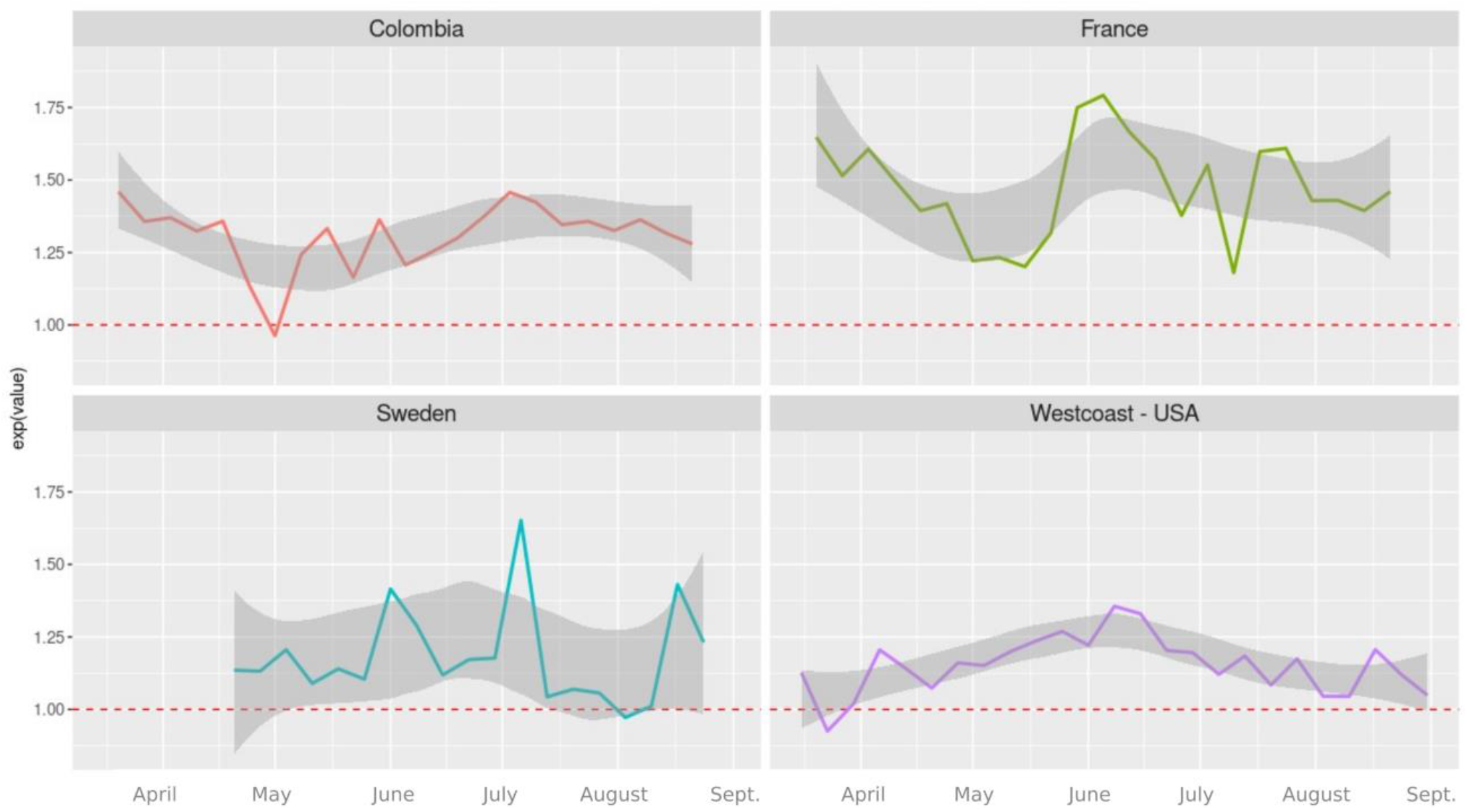
Multiplicative term on cases for log(FoI) variable unit incrementation, with 95% CI smoothing curves.

The interaction of the *log(FoI)* with the other main variable used, the number of cases registered in the previous week, was largely negative (Figure 6). This can be interpreted as follows: the influence of the *log(FoI)* in the regression was stronger in administrative units where fewer cases had been recorded the previous week. In other words, the impact of potentially imported cases is stronger in units that are lesser affected. This is particularly true at the very beginning of the epidemic in Colombia, which would underline the early role of mobility in the dissemination of the virus. This can be observed to a lesser but more continuous extent in France.

**Figure 6.**
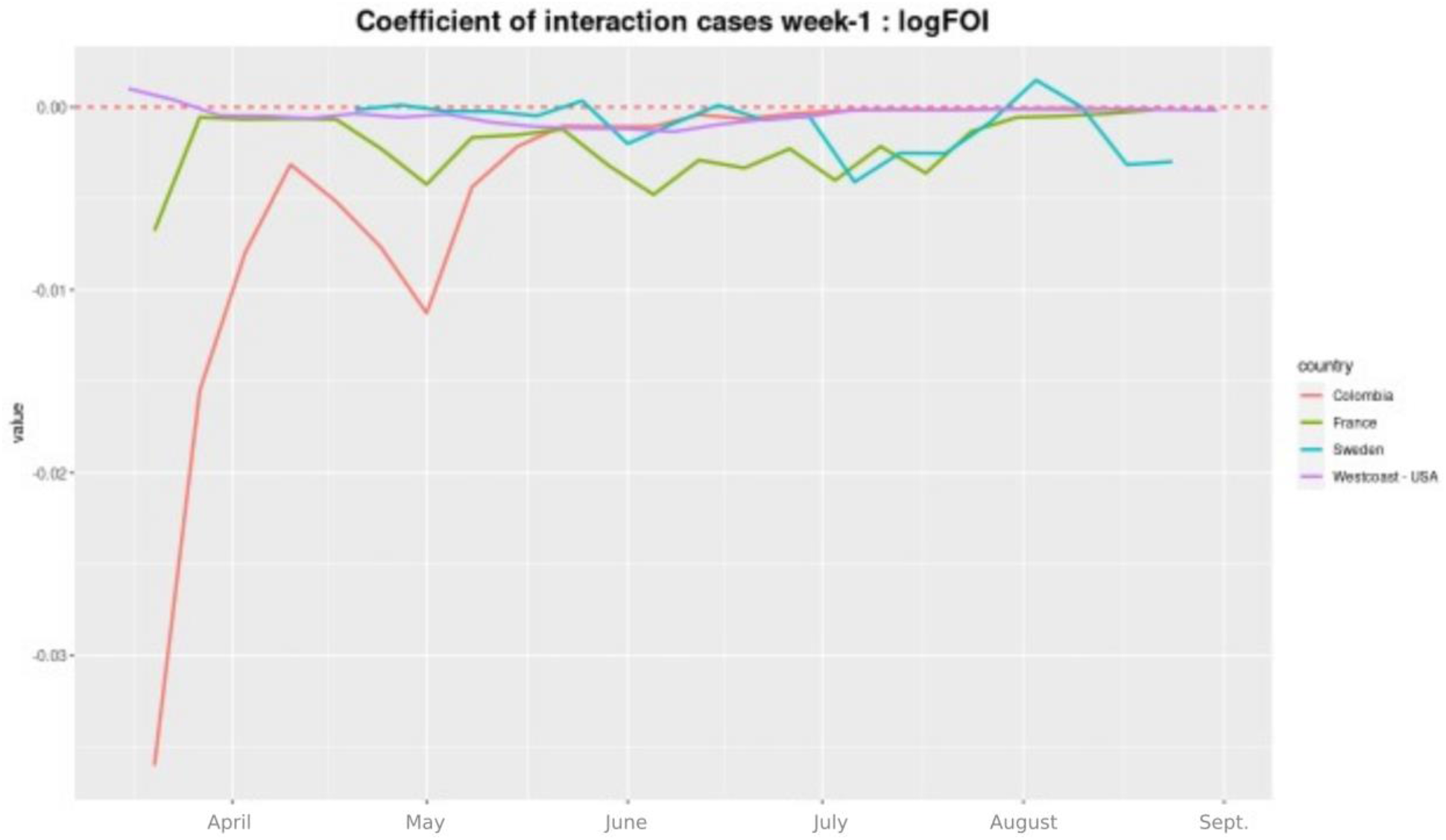
Coefficient of the interaction term between the log(FoI) and the number of cases of the previous week

Looking at the P-values of the *log(FoI)* variable (Figure 7) confirms this same tendency; this variable was consistently significant for both Colombia and France, exceeding P= 0.01 threshold very rarely. On average, P-values were generally below 0.05 in the USA but went beyond that threshold during 11 weeks (over a total of 25 weeks). The P-values in the Sweden models remained far superior to 0.05 for the vast majority of the time, suggesting that the link between *log(FoI)* and the cases of the following week was generally not significant.

**Figure 7.**
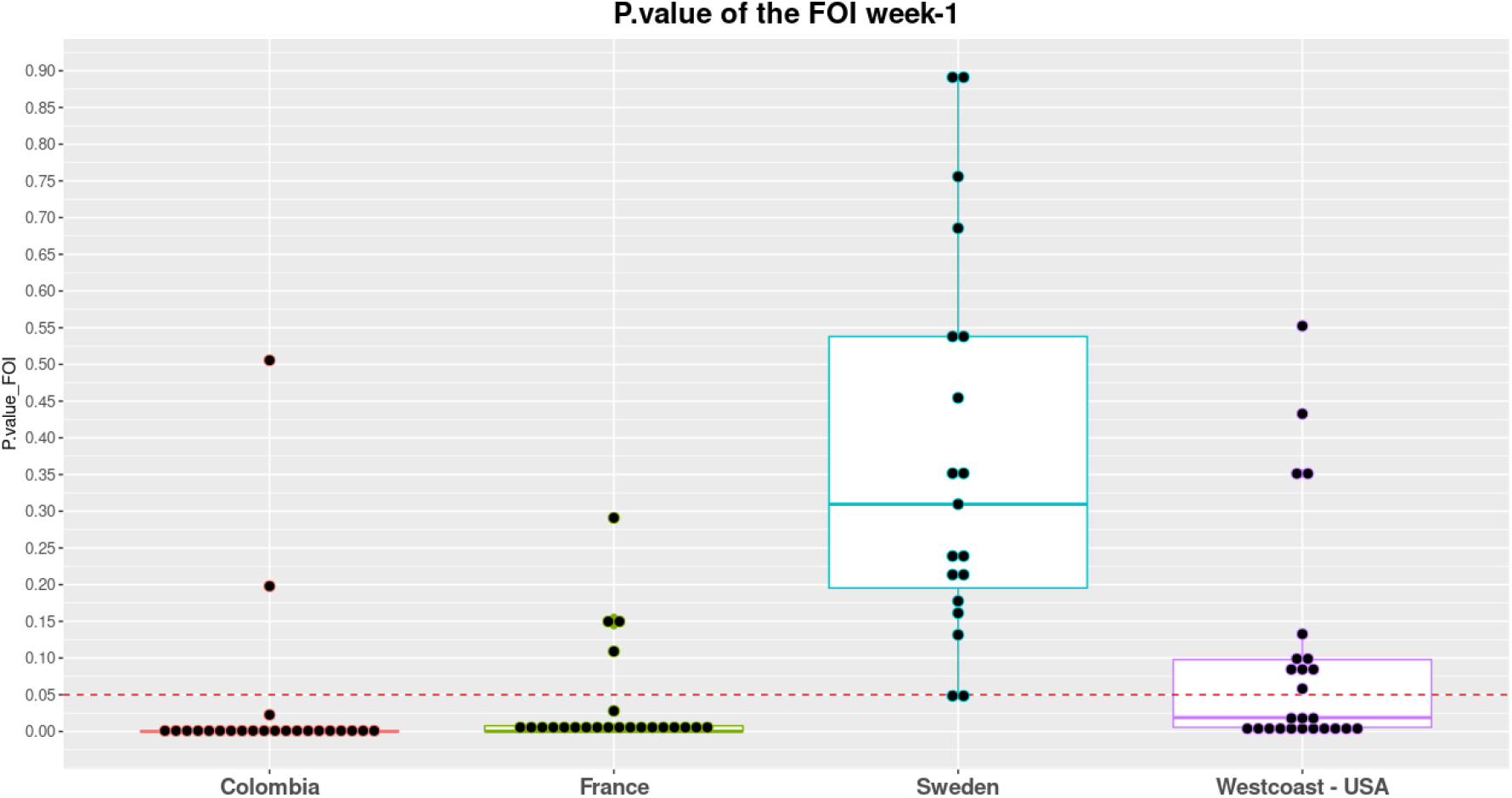
P values calculated for the log10(FoI) variable at each time step.

Figure 8 shows us a general assessment of the performance of the models through the calculation of McFadden’s Pseudo R^2^, which again reasserts that this predictive approach seems valid for Colombia and France, with median pseudo R^2^ of 0.68 and 0.60 respectively. For the USA, this indicator drops to 0.39 and for Sweden, to 0.33.

**Figure 8.**
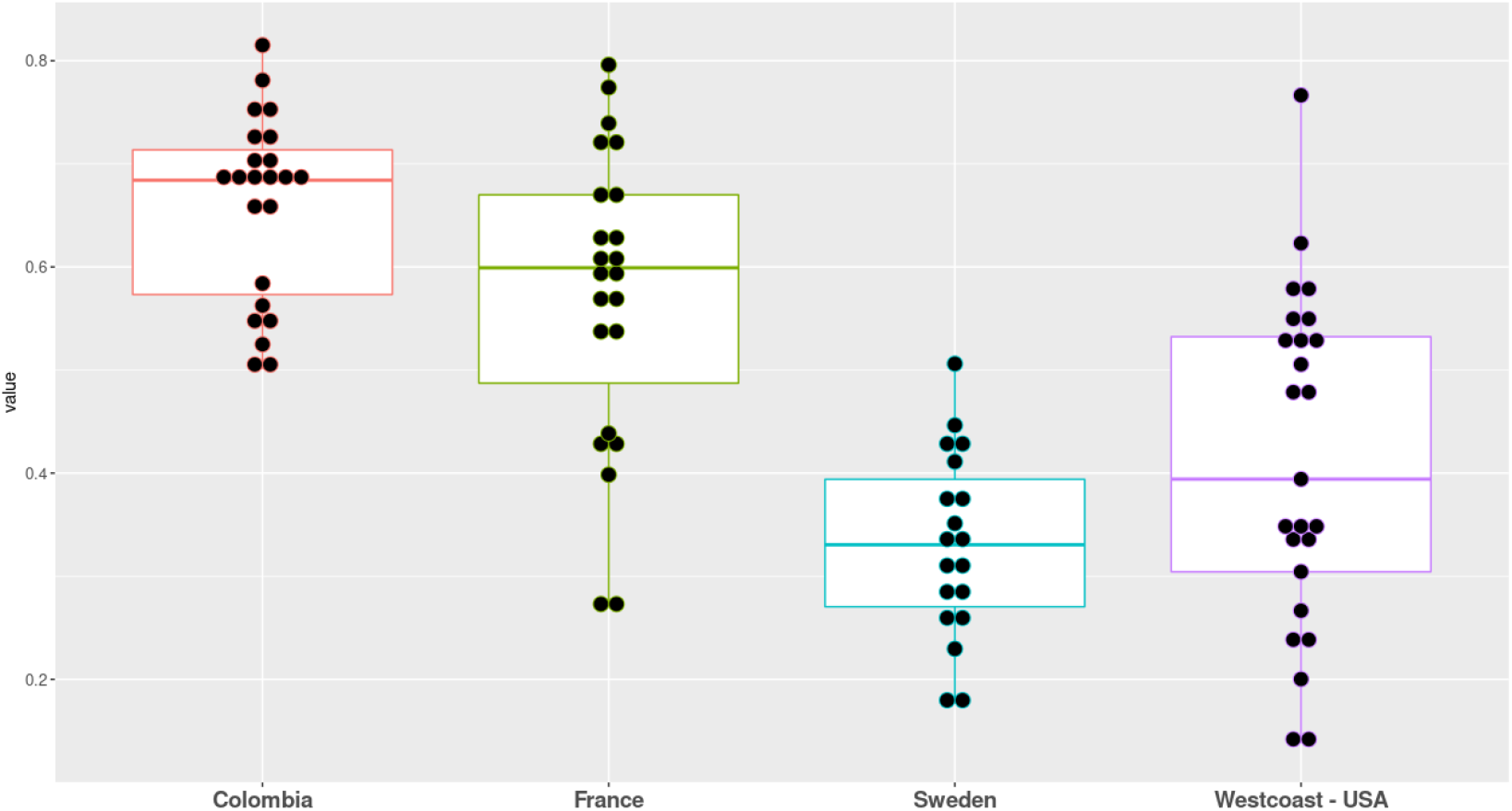
Goodness of fit measured by the McFadden’s Pseudo R2 of the respective models at each timestep

### Interpretation

Using the data collected by one of the major social media platforms, our analyses showed a great concentration of mobilities in metropolitan areas in non-lockdown periods. The three most important metropolitan areas concentrated more than 80% of inter-unit mobilities in Colombia, 79% in France, 61% in the West coast of USA and 50% in Sweden before the lockdown. These administrative units represent less than 10% of the overall number of units in each country. We also found that despite a drop in mobility in three of the four countries (USA, France, Colombia) just after the lockdown, the Gini index and the share of mobility captured by these metropolitan areas remained as high as prior to lockdown. Thus, whilst lockdown successfully reduced the mobility intensity in these countries, the mobility structure remained the same.

This phenomenon of human flow concentration can be problematic: just as a hyper connected individual can spread the virus disproportionately (a superspreader) (Lloyd-Smith et al. 2005; Kucharski et al. 2020; Lau et al. 2020) and large public gatherings lead to outbreaks (Ghinai et al. 2020; Hamner et al. 2020), a hyper-connected geographical unit can transmit the disease through its mobility network. Again, in three of the four countries studied, the Gini indices indicated an important concentration of cases in a small number of units; in each country, the most affected units were precisely those that were the most connected. The significant association of the mobility weighted force of infection and the much-improved model fit upon its inclusion in three of the four countries studied, supports the importance of considering contact intensity among units when analysing the risk of infection. This brings a paradigm change in the study of disease spread since it enables to apprehend the relational risk. The risk of Covid-19 in one administrative unit does not only depend on its intrinsic characteristics-such as human density or poverty-but is also a function of the epidemiological situation across the connectedness network.

If inter-individual contact is a fundamental element in the study of the spread of infectious diseases, it is also at the level of administrative units. However, this relational dimension is little understood beyond the individual scale. This can be imputed to the lack of data that can help to capture the mobility patterns in real-time. Until recently, mobility data have not been made available, were spatially aggregated and not provided in real-time. This is particularly an issue in a context of a lockdown since mobilities tend to change quickly and vary according to geographical scale. Fortunately, these types of data are getting increasingly provided by social media or mobile operators, and they can be used to help administrations to observe changes in mobility patterns and/or to better locate where to implement disease control operations (Pollina & Busvine 2020; Pullano et al. 2020; Romm et al. 2020).

The negative interaction between the previous week’s cases in the administrative unit of interest and the incoming force of infection into that unit suggests that as the association of one variable (*log10(FoI)* or previous week’s cases) on weekly cases becomes stronger, the other becomes weaker and vice versa. The very sharp decrease in the size of the (negative) coefficient in France and especially Colombia at the beginning of the study period mirrors almost exactly the decrease in the number of Facebook mobilities. This would suggest that the incoming *FoI*, at least at this early stage of the epidemic is not only having the major role in seeding infections, but that there is considerable heterogeneity among geographical units both with respect to numbers of cases and Facebook connectivities. That the interaction remains significant and negative throughout, despite the increasing number of cases, confirms the conclusions that this heterogeneity remains. This is confirmed by the Gini index that shows in every country a high concentration of both the cases and the mobility patterns.

Our analyses suggest a relatively modest impact of lockdown on the spread of the virus at the national scale since no country successfully implemented control measures to stem the spread of the virus despite varying levels of lockdowns. As observed in Hubei (Chinazzi et al. 2020), it is likely that the virus had already spread very widely prior to lockdown; the number of affected administrative units was already very high despite low testing levels. Only in Colombia was the initial number of affected units relatively low, but despite the strictest of lockdowns, affected units rose inexorably. Although undeniably important within a city, the contact intensity driven by hubs of connectivity clearly feed virus to bordering locations and this flow is maintained despite drastic reductions in mobilities at least over longer distances. Once seeded, the virus then spreads among locally connected units. However, the fact that the FoI is more significant in countries who implemented a drastic lockdown suggests that mobility reduction helps to predict paths of diffusion according to remaining mobilities. While we underlined that this mobility remained high mostly in metropolitan areas during the lockdown, it suggests that disease control implemented in these areas could have a maximum impact over the short term. This element, combined with the interaction between the FOI and the previous weeks cases is particularly promising: it suggests that vaccine implementation in the most connected units of a highly affected area could prevent the geographical spread of the virus.

In constructing a framework to integrate mobility patterns into an understanding of pathogen diffusion to guide the way diseases are managed in a long-term perspective, the question arises of properly managing diseases in a context of exacerbated mobility that cannot be completely stopped at the local scale. As such, the redefinition of the systems depending of connectivity between units should make it possible to better manage epidemics by aligning the area of action of health policies with the area of dissemination of infectious diseases. However, as they are currently conceived, health policies and infrastructures are very largely circumscribed according to the administrative boundaries of municipalities / regions, which no longer correspond to the realities of daily mobility.

### Limitations

This study is clearly limited in two aspects: the quality of the case data and the non-representativity of Facebook users and the general population. However, this being said, it is remarkable that such a simple combined Facebook infection measure yields consistent results across three very different settings. Facebook users tend to be biased to the adult population, who are in turn more likely to travel between administrative units for work purposes and thus more likely to interact with other adults. Insofar as the epidemiological role of children is now believed to be significantly less than that of adults, the Facebook data may actually give a more representative picture of the flow of the important epidemiological sector of the population.

In conclusion, human mobility has played a significant role in the spread of SARS-CoV-2 and to an extent that could have been predicted from the mobility network. Incorporation of such network information would significantly aid in developing more refined public health strategies for disease management and control both in the short and long term.

## Data Availability

All the data are fully available on request

https://dataforgood.fb.com/

https://www.santepubliquefrance.fr/

https://www.cdc.gov/

https://www.minsalud.gov.co/Paginas/default.aspx

https://www.folkhalsomyndigheten.se/the-public-health-agency-of-sweden/

---

**Supplementary Figure 1:**
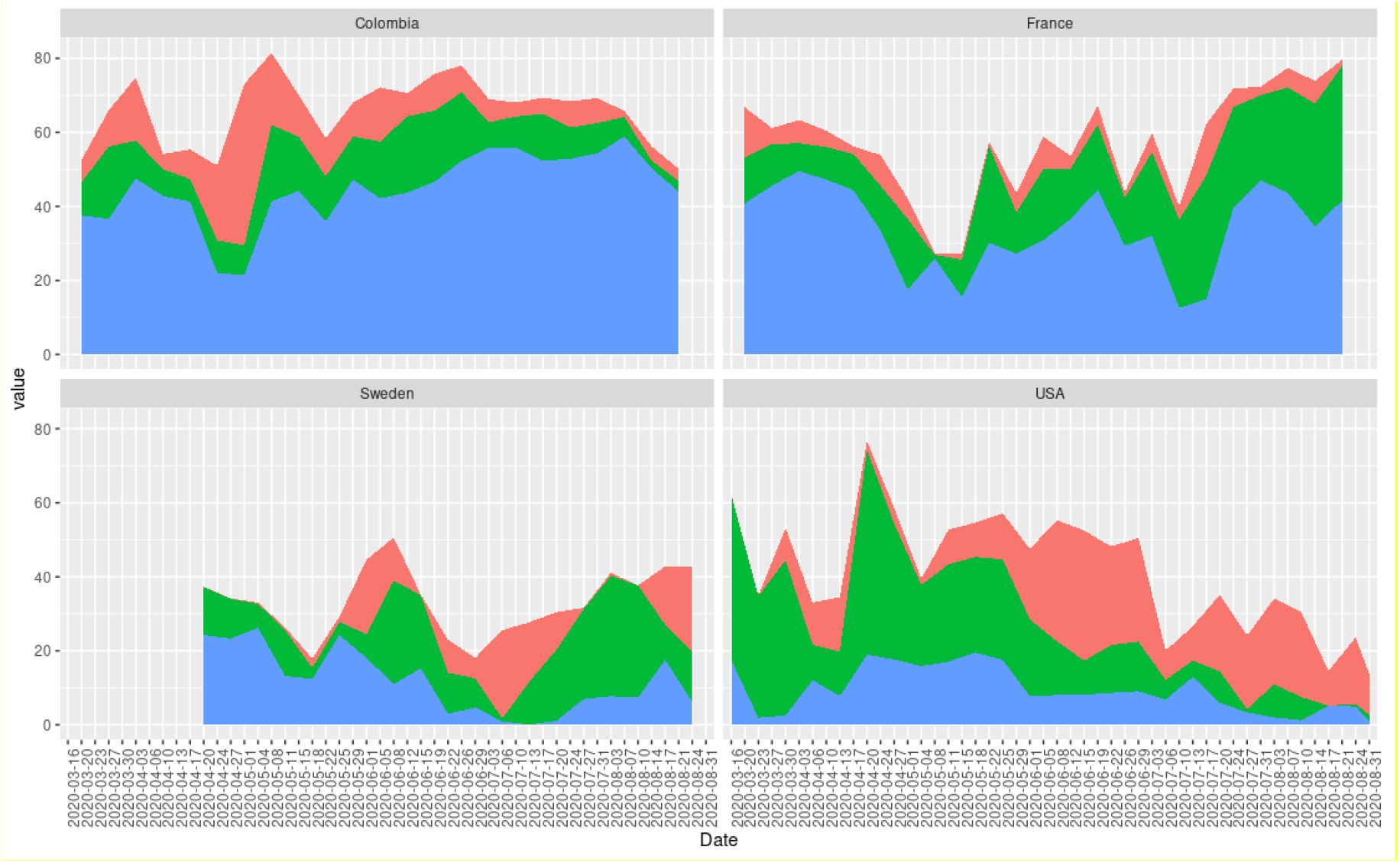
Cumulative percentage of deviance explained by each variable (Blue-FoI; Green – cases the previous week; Pink – the interaction between FoI and previous week’s cases).

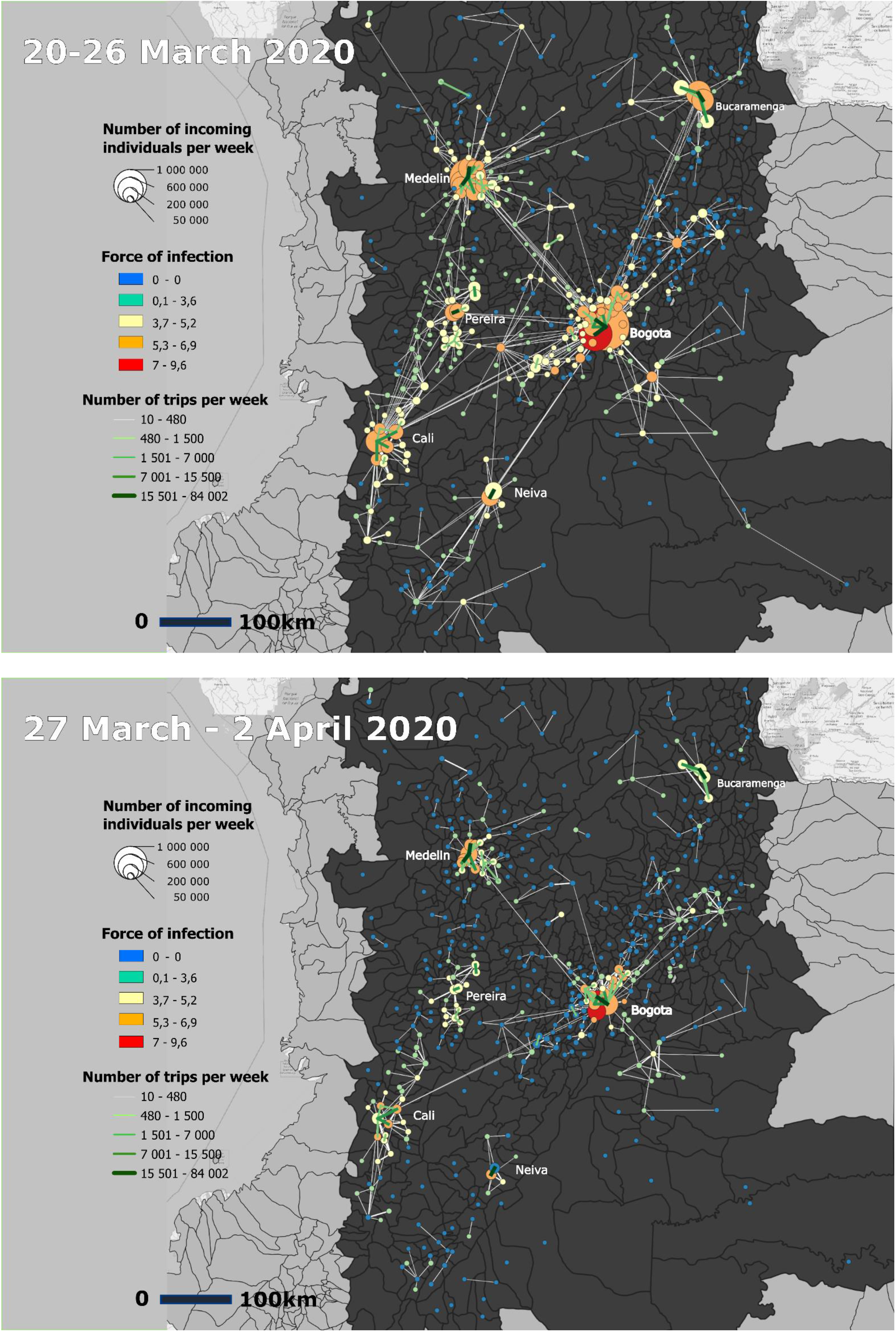
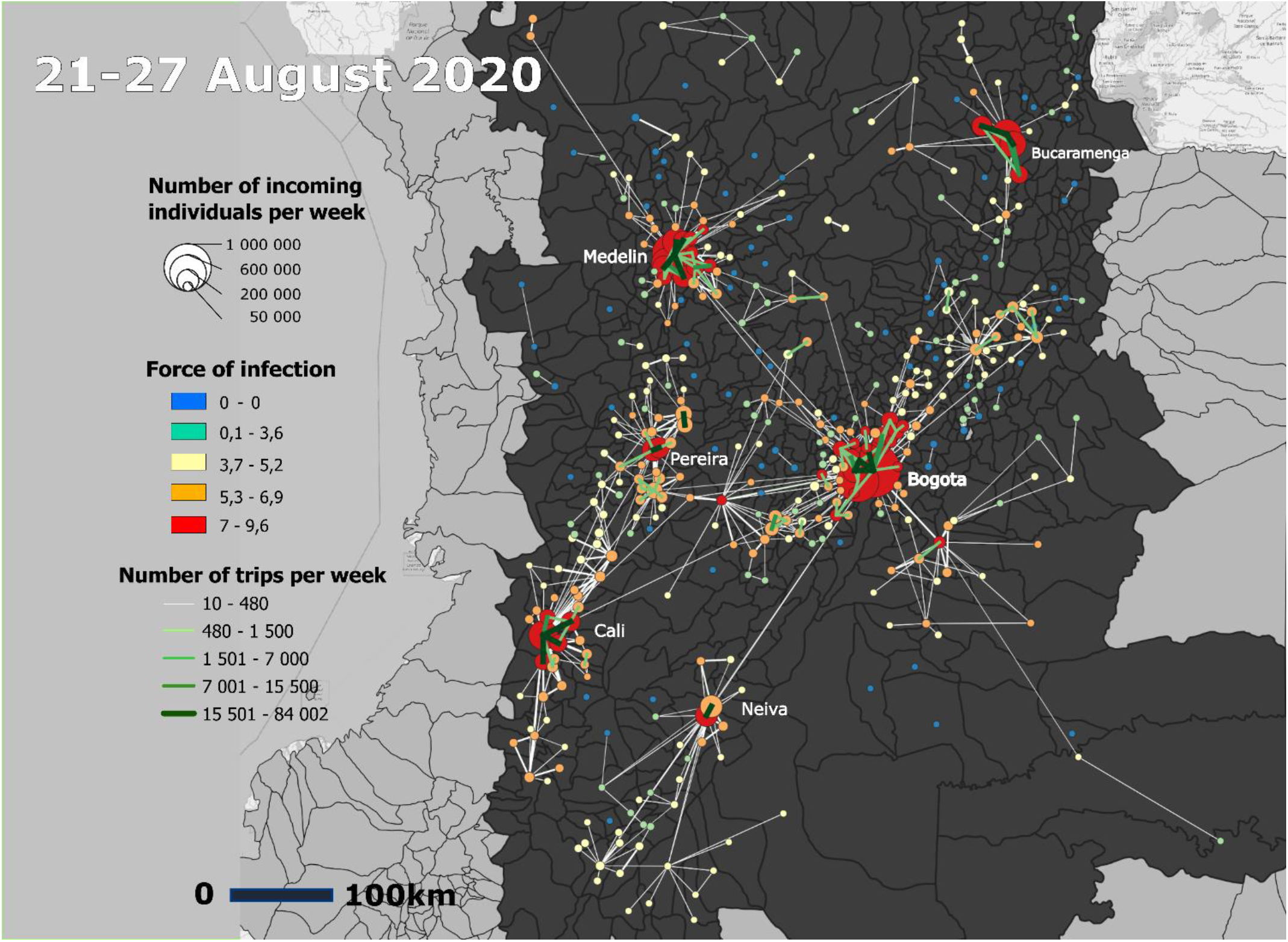

Collection of maps 1 : Mobility flux, number of incoming mobility and FOI Before the lockdown, during the lockdown and in August in Colombia

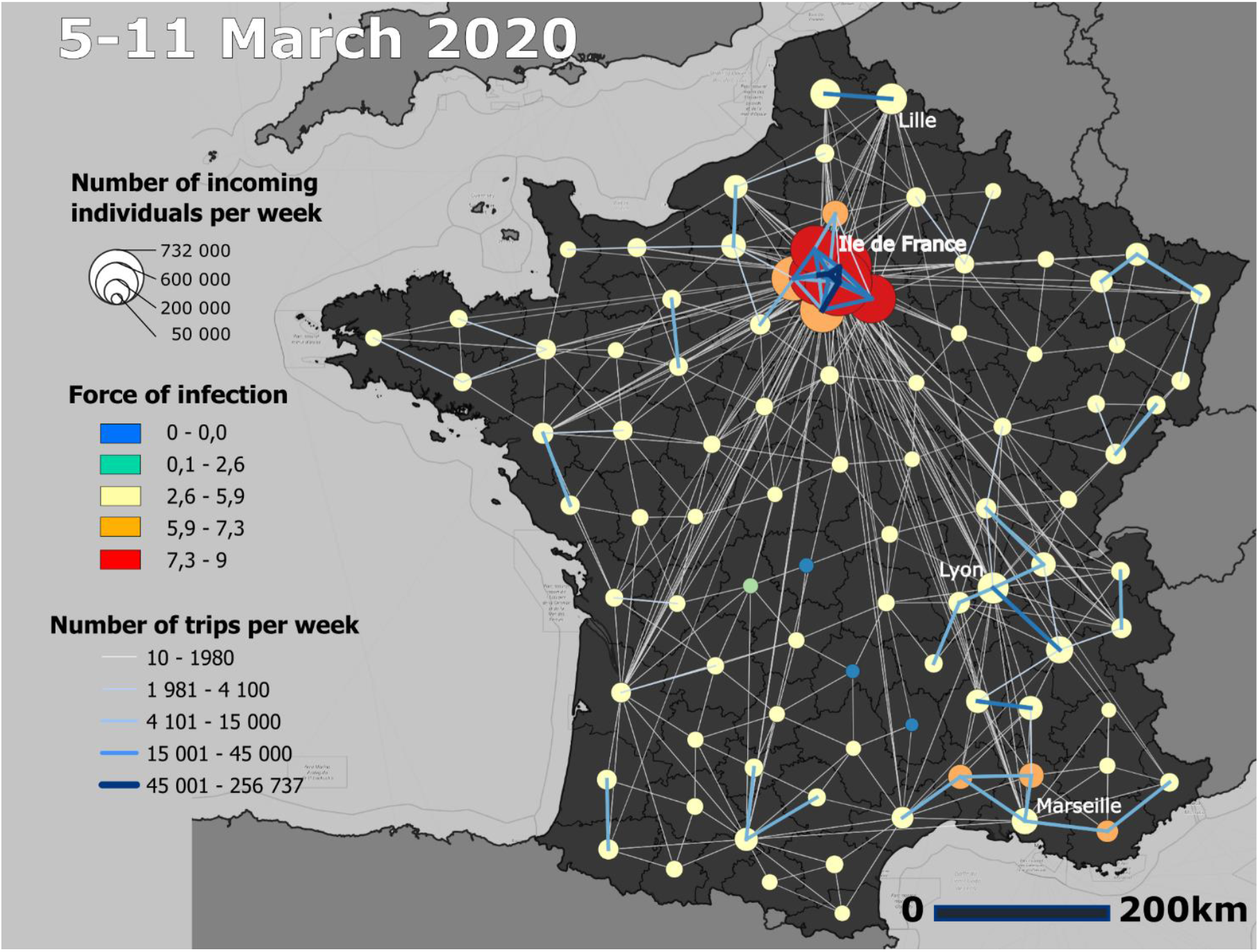
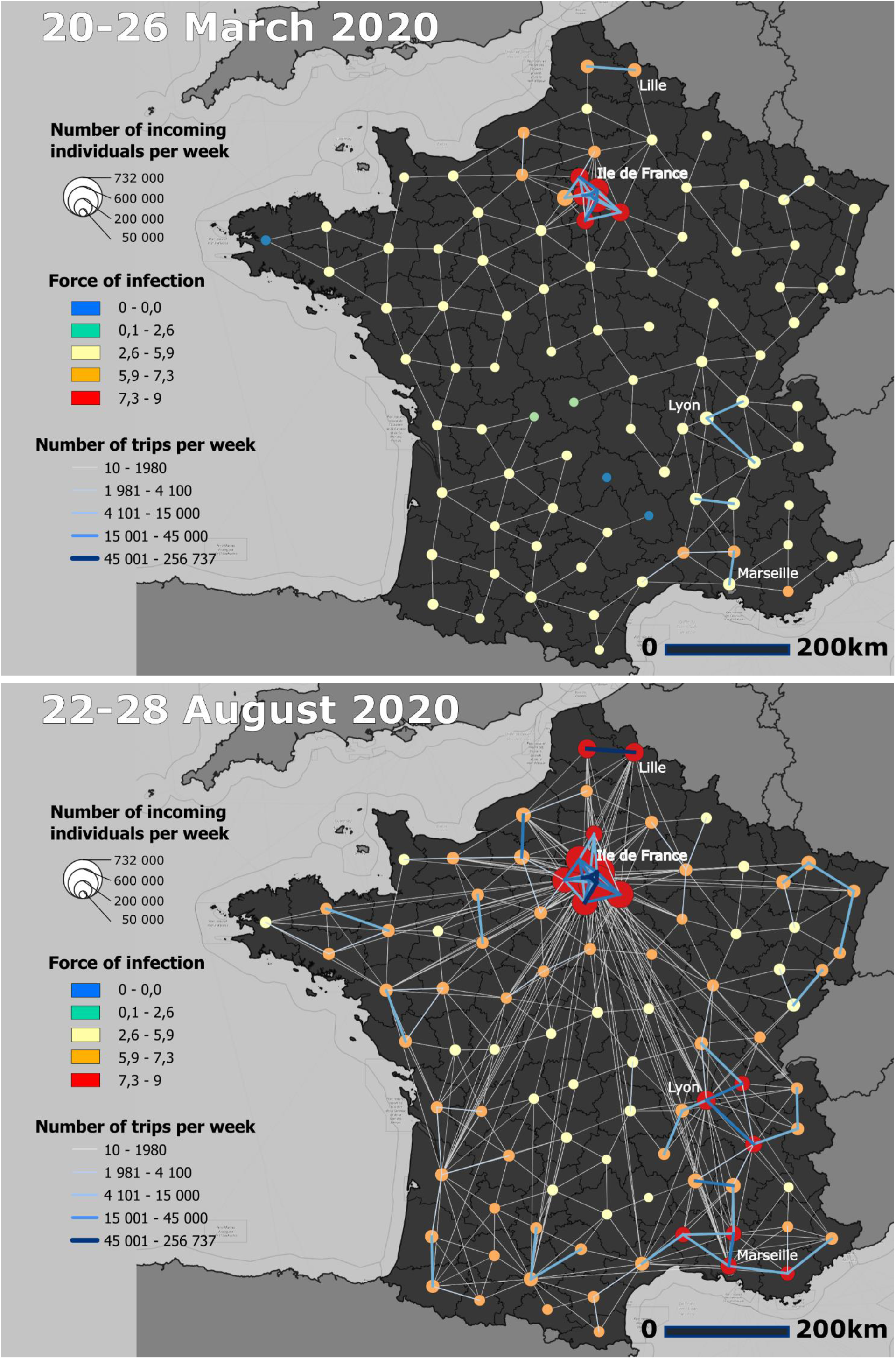

*Collection of maps 2* : *Mobility flux, number of incoming mobility and FOI Before the lockdown, during the lockdown and in August in France*

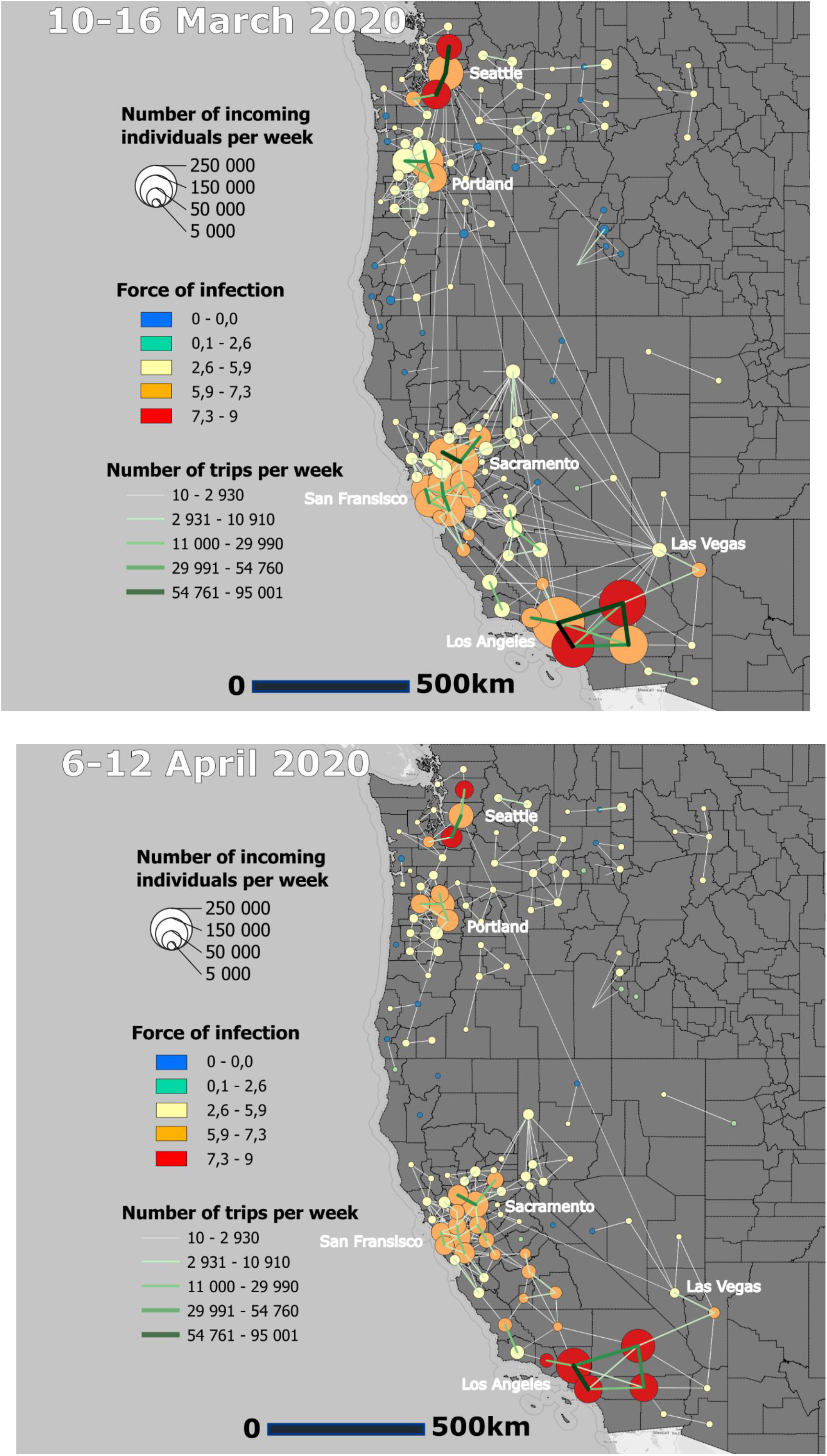
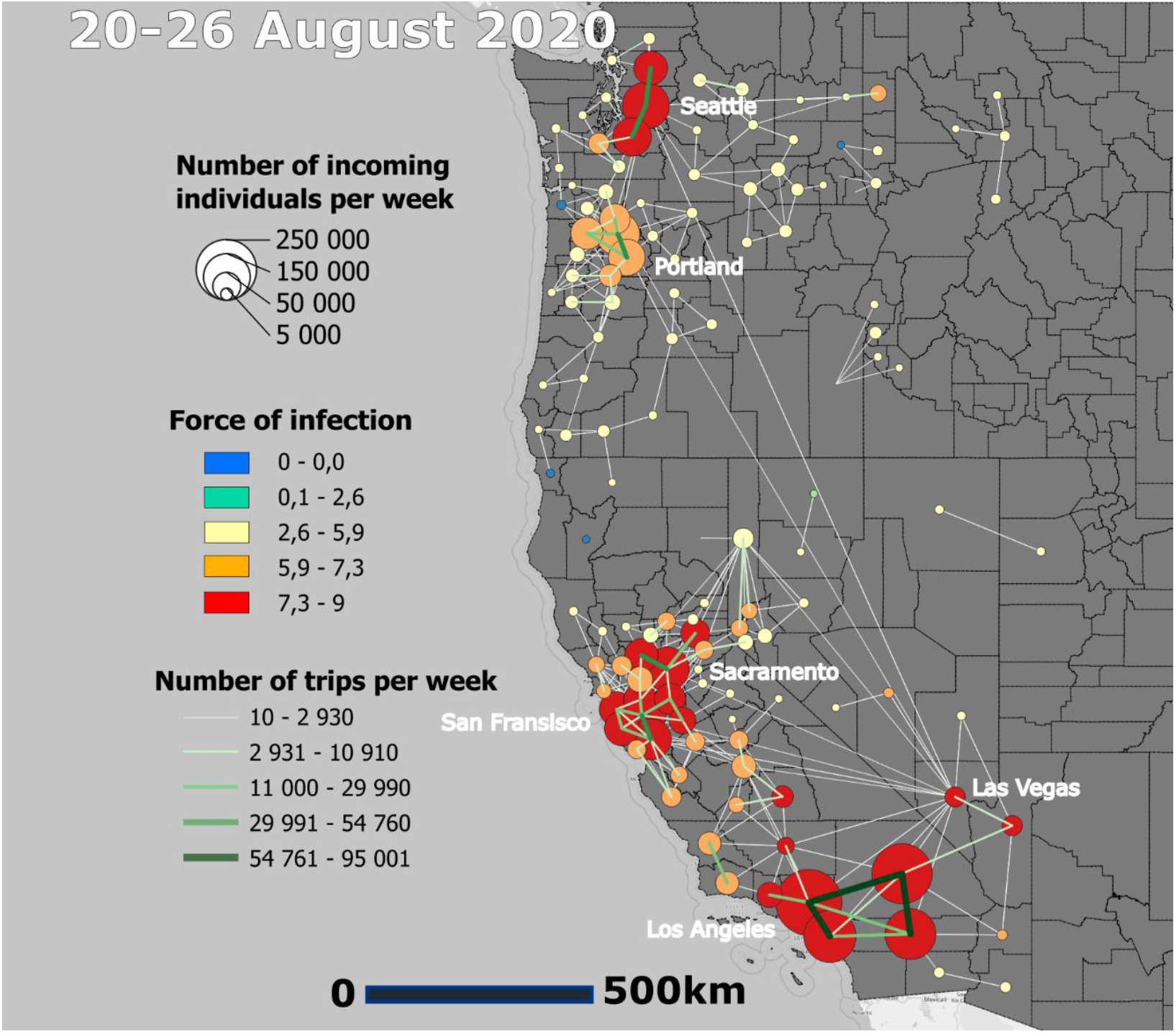

*Collection of maps 3 : Mobility flux, number of incoming mobility and FOI Before the lockdown, during the lockdown and in August in the west coast of USA*

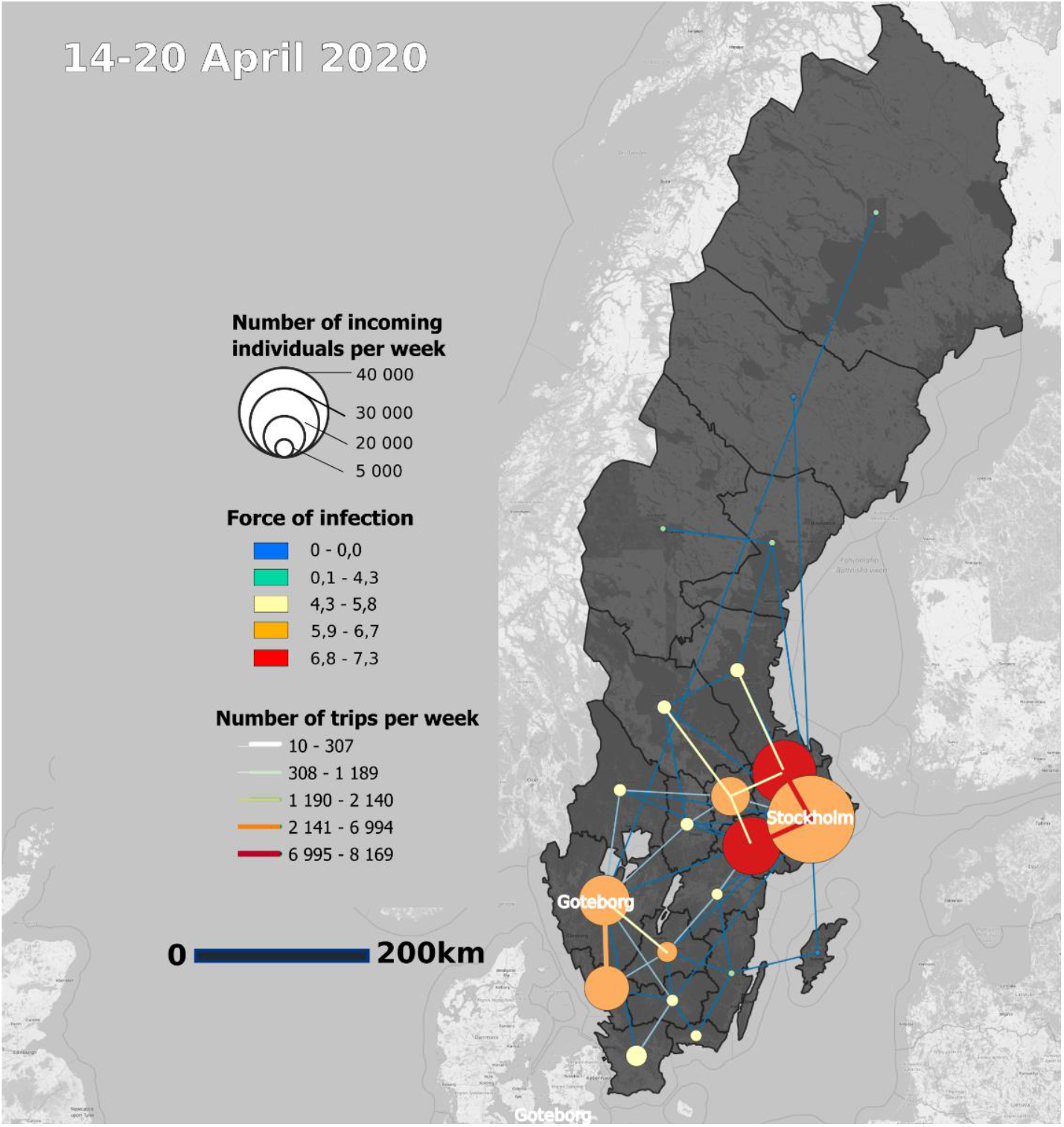
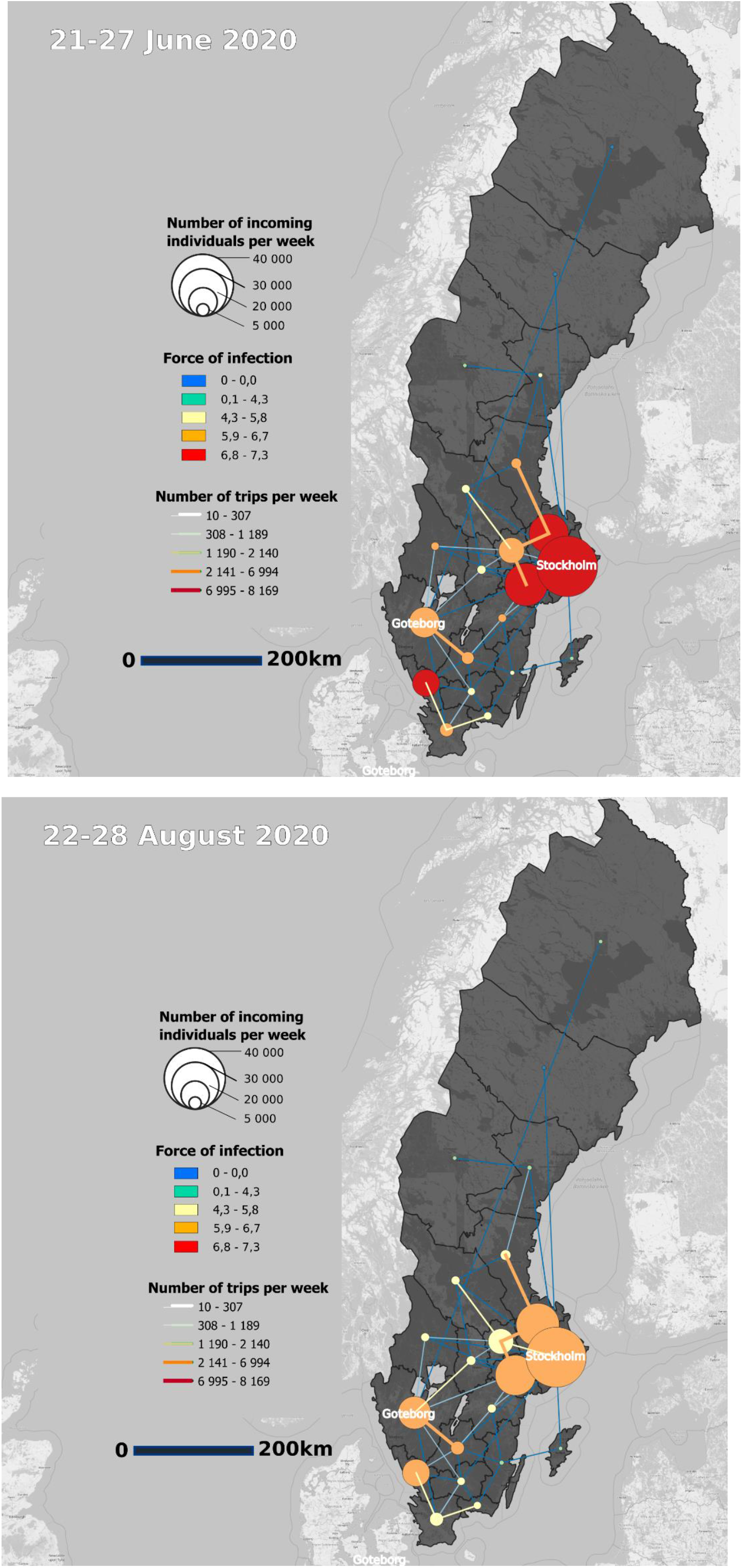

*Collection of maps 4: Mobility flux, number of incoming mobility and FOI In April, June and in August in Sweden*

